# Blood-brain barrier dysfunction in cerebral amyloid angiopathy is associated with disseminated cortical superficial siderosis

**DOI:** 10.64898/2026.06.17.26355799

**Authors:** Berkant Bay, Malte Pfister, Hendrik Mattern, Jose Bernal, Katja Neumann, Marc Dörner, Sven G. Meuth, Stefanie Schreiber, Philipp Arndt

**Author notes:** **Corresponding author:** Philipp Arndt, Otto-von-Guericke University Magdeburg Leipziger Str. 44, 39120 Magdeburg, Germany.

## Abstract

**Background:** Blood–brain barrier (BBB) dysfunction is increasingly recognized as a feature of cerebral amyloid angiopathy (CAA) and has been linked to hemorrhagic imaging manifestations such as cortical superficial siderosis. However, it remains unclear whether neurovascular barrier dysfunction can be captured by routinely available fluid biomarkers and whether such markers identify clinically relevant hemorrhage-prone CAA phenotypes. The CSF/serum albumin quotient (QAlb) is an established marker of neurovascular barrier dysfunction. We investigated Q_Alb_ levels in CAA and their association with imaging markers of disease severity.

**Methods:** We included 225 participants (115 with CAA, 72 with Alzheimer’s disease [AD], 38 healthy controls) with CSF biomarkers and standardized MRI evaluation. Pathologic Q_Alb_ levels were identified via the age-corrected Reiber-formula. Group differences and determinants of pathological Q_Alb_ were assessed using uni- and multivariable regression analyses. The diagnostic relevance was assessed by receiver operating characteristic analysis.

**Results:** Q_Alb_ levels were higher in CAA than in controls (ratio of means [RoM] 1.43, 95% CI 1.28–1.58) and patients with AD (RoM 1.22, 95% CI 1.10–1.35; both p<0.001). Pathological Q_Alb_ was independently associated with CAA compared with controls (OR 12.16, 95% CI 2.56–57.86) and AD (OR 2.14, 95% CI 1.07–4.28). Despite these associations, Q_Alb_ showed only moderate discrimination between CAA and controls (AUC 0.75, 95% CI 0.67–0.82) and low discrimination between CAA and AD (AUC 0.63, 95% CI 0.55–0.71). Within the CAA cohort, pathological Q_Alb_ was independently associated with disseminated cortical superficial siderosis (OR 3.92, 95% CI 1.31–11.72; p=0.014), a marker of advanced hemorrhage-prone disease.

**Conclusion:** QAlb is elevated in patients with CAA and is associated with disseminated cortical superficial siderosis, the strongest predictor of future intracerebral hemorrhage. These findings support an association between neurovascular barrier dysfunction and the hemorrhage-prone phenotype of CAA.

## Background

Cerebral amyloid angiopathy (CAA) is a common age-related small vessel disease and a major cause of lobar intracerebral hemorrhage (ICH) and cognitive impairment in older adults [1, 2]. The neuropathological hallmark of CAA is the progressive deposition of β-amyloid (Aβ) in the walls of cortical and leptomeningeal arterioles. Beyond vascular Aβ deposition, growing neuropathological and neuroimaging evidence suggests that blood–brain barrier (BBB) dysfunction and vascular inflammation contribute to disease progression. Emerging neuroimaging evidence suggests that, beyond amyloid deposition, vascular inflammation and blood–brain barrier (BBB) dysfunction may contribute to disease progression [3–5]. Human proteomic and neuropathological studies further support inflammatory and immune-mediated mechanisms in sporadic CAA, including barrier dysfunction, complement activation and perivascular inflammatory remodeling [6, 7]. However, it remains unclear whether neurovascular barrier dysfunction represents a disease-specific feature of CAA rather than a nonspecific consequence of ageing or neurodegeneration, and whether it is linked to hemorrhage-prone manifestations of the disease.

Recent MRI studies have demonstrated BBB leakage *in vivo* in patients with CAA and linked leakage to cortical superficial siderosis (cSS), a marker of advanced leptomeningeal vessel injury and future ICH risk [5]. However, it is unknown whether these abnormalities can be captured by routinely available fluid biomarkers and whether such markers identify clinically relevant CAA phenotypes. The CSF/serum albumin quotient (Q_Alb_) is an established marker of neurovascular barrier dysfunction. We therefore investigated whether Q_Alb_ is elevated in patients with CAA compared with Alzheimer disease (AD) patients and controls without imaging evidence of CAA, and whether pathological Q_Alb_ levels are associated with MRI markers of advanced hemorrhage-prone CAA.

## Methods

### Study population

We included 225 participants, comprising n=115 patients with probable CAA, n=72 patients with cognitive impairment due to AD and n=38 cognitively unimpaired controls. Inclusion criteria were: (i) MRI with iron-sensitive sequences, and (ii) lumbar puncture for CSF collection and analysis during diagnostic workup for all participants. Patients with probable CAA were selected from our prospectively curated database on CAA between February 2011 and October 2025 at the Department of Neurology, Otto-von-Guericke University Magdeburg, as recently described [8]. Probable CAA was based on the Boston criteria version 2.0 [9]. AD patients and controls were selected from our institutional memory clinic database. AD patients had mild cognitive impairment or dementia and positive CSF biomarkers suggestive of AD pathology (A+T+), according to the NINCDS/ADRDA and ATN criteria [10, 11]. Controls were elderly patients who also presented to the memory clinic but showed no cognitive deficits on comprehensive neuropsychological testing. All controls and AD patients were required to have no hemorrhagic markers on MRI, i.e. no ICH, convexity subarachnoid hemorrhage (cSAH), cerebral microbleeds (MB) and no cortical superficial siderosis (cSS) and were recruited between February 2015 and May 2025. AD patients and controls were identified through a positive selection process, including all individuals meeting the predefined diagnostic and imaging criteria during the recruitment period.

### Clinical data and neuropsychological assessment

Patients were characterized with regard to demographics (age, sex), clinical phenotype at presentation (cognitive impairment, hemorrhagic onset, i.e. lobar ICH or cSAH, transient focal neurological episodes, history of ischemic stroke, history of seizure or a mixture of them), and vascular risk factors, i.e. arterial hypertension, type 2 diabetes and hyperlipidemia based on former diagnosis on medical records and/or use of antihypertensive, antidiabetic or lipid lowering medication.

### Cerebrospinal fluid biomarkers

CSF samples were centrifuged at 4 °C, aliquoted and stored at −80 °C until analysis. CSF biomarker levels were available for markers of AD pathology (Aβ_42/40_ ratio, pTau_181_). AD pathology biomarker levels were determined with ELISA kits (until 11/2019: Innotest Aβ_40_, Innotest Aβ_42_, Innotest pTau, Innogenetics, Ghent, Belgium) or automated immunoassays (from 12/2019: LUMIPULSE® G600 II, Fujirebio Inc., Japan). Each participant was categorized as having normal AD biomarker levels (A-T-), AD pathological changes (A+T-) or AD pathology (A+T+). “A” was based on the Aβ_42/40_ ratio ([Aβ_42_ / Aβ_40_] x 10) and “T” on pTau levels. The following locally established thresholds were applied for this classification: using ELISA 0.50 for Aβ_42/40_ ratio and 70 pg/mL for pTau, and using automated immunoassays 0.69 for Aβ_42/40_ ratio and 56 pg/mL for pTau. Q_Alb_ assessed BBB dysfunction. CSF and serum albumin levels were analyzed by rate nephelometry (Immage 800, Beckman Coulter) [12].

### MRI acquisition and analysis

For analysis, clinical 3T MRI (44.4%, n=34 Controls, n=53 CAA, n=13 AD) or 1.5T MRI (55.6%, n=4 Controls, n=62 CAA, n=59 AD; both Siemens Healthineers, Erlangen, Germany) was used to quantify ICH, cSAH, MB, cSS, white matter hyperintensities (WMH) and lacunes according to the Standards for Reporting Vascular Changes on Neuroimaging (STRIVE) criteria by one trained investigator (M.P.) blinded to CSF and clinical data [13, 14]. The following sequences were used: T2*-weighted gradient-recalled echo for MB, cSS, and ICH, T2-weighted for perivascular spaces (PVS) in the basal ganglia (BG) and centrum semiovale (CSO) and T2-weighted fluid-attenuated inversion recovery (FLAIR) for white matter hyperintensities (WMH) and lacunes. Presence and number of MB, ICH or lacunes were assessed according to the Microbleed Anatomic Rating Scale and the Cerebral Hemorrhage Anatomical RaTing instrument [15, 16]. WMH in deep and periventricular regions were rated according to the Fazekas scale and specific WMH patterns as recently proposed [17, 18].

### Statistical analysis

Results of variables were expressed as median (interquartile range [IQR]), mean (standard devidation [(SD]), or proportions, as appropriate. Intergroup comparisons of demographic variables, vascular risk factors, and Q_Alb_ levels between clinical groups (CAA vs. AD vs. controls) were performed in univariate analyses, using the χ^2^-test, Kruskal-Wallis test or ANOVA, as appropriate. Pairwise post-hoc comparisons were conducted applying Kruskal-Wallis-test with Dunn’s correction, χ^2^-test and ANOVA with Bonferroni’s correction to control for multiple testing.

For Q_Alb_, we used the ratio of means (RoM) as an effect measure, calculated as Q_Alb_ level ratio between groups of interest. This method controls variability between studies and allows to integrate the marker into the existing core CSF biomarker profile of CAA patients. A ratio above 1 indicates that the concentration of the biomarker is higher in CAA vs. the comparison population, and a ratio below 1 indicates the concentration is higher in the comparison group. Standard errors of the RoM CSF values were calculated using the delta method, and mean values are presented with 95% confidence intervals (CI) [19, 20]. Further, pathologic Q_Alb_ levels were identified via the age-corrected Reiber-formula (pathologic Q_Alb_ threshold = 4 + [Age in years / 15]) [21]. Independent associations were tested in multivariate linear and logistic regression models, adjusted for biologically relevant confounding variables. Receiver operating characteristic (ROC) analysis was performed to assess the discriminative ability of Qalb levels, with optimal cut-off values determined by maximizing Youden’s J statistic. Sensitivity and specificity values for Q_Alb_ reflect performance at these thresholds. To investigate which variables drive increased Q_Alb_ levels in CAA, patients were grouped according to pathologic Q_Alb_ levels. Variables of interest were demographics, clinical presentation, neuroimaging markers, and neurodegenerative co-pathologies. Intergroup comparisons were performed in univariate analyses, using the χ^2^-test, t-test or Mann-Whitney U-test, as appropriate. Significant variables were entered into a logistic regression analysis to assess independent associations. Significance level was set at p<0.05 for all analyses. IBM SPSS Statistics 24.0 software was used for all statistical analyses.

## Results

We included 225 participants with a median age of 76 years (IQR 71–81), of whom 103 (45.8%) were female. Of those, 115 patients had probable CAA according to the Boston criteria version 2.0, 72 patients had cognitive impairment due to AD and 38 were healthy, cognitively unimpaired elderly controls. Controls were significantly younger than patients with CAA and AD and had less frequent history of arterial hypertension and type 2 diabetes. AD patients were more frequently female than the remainder. Key characteristics of each subgroup are shown in **Table 1**.

**Table 1.** Key characteristics of each patient subgroup. Pathologic Q_Alb_ levels were identified via the age-corrected Reiber-formula (4 + [Age in years / 15]). Abbreviations: AD, Alzheimers Disease; CAA, cerebral amyloid angiopathy: CON, controls.

|  | CON<br>n = 38 | CAA<br>n = 115 | AD<br>n = 72 | p-value |
| --- | --- | --- | --- | --- |
| Age | 72 (63–74) | 76 (71–81) | 75 (71–79) | **p < 0.001** |
| Female sex | 15 (39.5%) | 46 (40.0%) | 42 (58.3%) | **p = 0.035** |
| Hypertension | 21 (55.3%) | 99 (86.1%) | 55 (76.4%) | **p < 0.001** |
| Dyslipidemia | 13 (34.2%) | 43 (37.4%) | 23 (31.9%) | p = 0.74 |
| Type 2 Diabetes | 1 (2.6%) | 36 (31.3%) | 22 (30.6%) | **p = 0.001** |
| $Q_{Alb}$ | 5.99 (5.12–7.19) | 7.99 (6.03–10.92) | 7.05 (5.45–8.37) | **p < 0.001** |
| Pathologic $Q_{Alb}$ | 2 (5.3%) | 46 (40.0%) | 16 (22.2%) | **p < 0.001** |

### Q_Alb_ is elevated in CAA patients compared to AD patients and controls

Individuals with CAA had significantly higher Q_Alb_ levels compared to controls (RoM 1.43, 95% CI 1.28–1.58; median [IQR] CAA 7.99 [6.03–10.92] vs. controls 5.99 [5.12–7.19], p<0.001] and those with AD (RoM 1.22, 95% CI 1.10–1.35; CAA 7.99 [6.03–10.92] vs. AD 7.05 [5.45–8.37], p=0.010]. Findings were consistent even when restricting analyses to CAA patients fulfilling Boston criteria v1.5 (n=98, 85.2%) with a more hemorrhagic phenotype (for controls: RoM 1.42, 95% CI 1.24–1.61; CAA 7.90 [6.02–11.01] vs. controls 5.99 [5.12–7.19], p<0.001; for AD: RoM 1.22, 95% CI 1.06–1.37; CAA 7.90 [6.02–11.01] vs. AD 7.05 [5.45–8.37], p=0.029]. Individuals with a diagnosis of CAA were more likely to have pathologically elevated Q_Alb_ levels than controls (OR = 12.16, 95% CI 2.56–57.86, p = 0.002) and patients with AD (OR = 2.14, 95% CI 1.07–4.28, p = 0.031; **Table 2**), even after adjustment for demographic variables and vascular risk factors.

**Table 2.** Independent associations between CAA diagnosis and pathologic Q_Alb_ (Reiber). Logistic regression analysis with Pathologic Q_Alb_ as the dependent variable was performed to determine whether demographic variables and vascular risk factors influence the association with CAA diagnosis, between (i) CAA and controls and (ii) between CAA and AD. Pathologic Q_Alb_ levels were identified via the age-corrected Reiber-formula (4 + [Age in years / 15]). Regression coefficients (B) with 95% confidence intervals (CI) and p-values are shown. Abbreviations: AD, Alzheimer’s disease; CAA, cerebral amyloid angiopathy.

| Comparison | Dependent variable | Independent variables | OR (95%CI) | p-value |
| --- | --- | --- | --- | --- |
| CAA versus controls | Pathologic $Q_{Alb}$ (Reiber) | CAA diagnosis | 12.16 (2.56–57.86) | **0.002** |
|  |  | Age | 1.00 (0.95–1.06) | 0.89 |
|  |  | Female sex | 0.43 (0.19–0.97) | **0.041** |
|  |  | Hypertension | 0.72 (0.25–2.12) | 0.55 |
|  |  | Type 2 Diabetes | 1.54 (0.66–3.58) | 0.32 |
| CAA versus AD | Pathologic $Q_{Alb}$ (Reiber) | CAA diagnosis | 2.14 (1.07–4.28) | **0.031** |
|  |  | Age | 1.00 (0.95–1.05) | 0.99 |
|  |  | Female sex | 0.53 (0.27–1.03) | 0.06 |
|  |  | Hypertension | 0.91 (0.37–2.23) | 0.84 |
|  |  | Type 2 Diabetes | 1.53 (0.77–3.06) | 0.23 |

### Q_Alb_ has low to moderate diagnostic potential to detect CAA patients

Q_Alb_ had moderate discrimination between CAA and controls (AUC 0.75 [95% CI 0.67–0.82], sensitivity 57.4%, specificity 92.1%, cutoff 7.64), and low discrimination between CAA and AD (AUC 0.63 [95% CI 0.55–0.71], sensitivity 59.1%, specificity 65.3%, cutoff 7.53). The accuracy did not change considering only CAA patients according to the Boston criteria version 1.5 (for controls: AUC 0.74 [95% CI 0.65–0.82], sensitivity 55.1%, specificity 92.1%, cutoff 7.64; for AD: AUC 0.62 [95% CI 0.54– 0.70], sensitivity 57.1%, specificity 65.3%, cutoff 7.53).

### Predictors of pathologic Q_Alb_ within CAA

Next, we investigated whether specific demographic, clinical, neuroimaging, or CSF features were associated with pathologic Q_Alb_ levels within CAA patients. CAA patients with pathologic Q_Alb_ (Q_Alb_ range 8.20–19.17) were compared to the rest of the CAA group (Q_Alb_ range 2.89–8.81, **Table 3**). Pathologic Q_Alb_ levels were found less frequently in female CAA patients (OR 0.42 [95% CI 0.18–0.95]; p=0.038) and more frequently in those with disseminated cSS (OR 3.92 [95% CI 1.31–11.72], p=0.014, **Table 4**).

**Table 3.** Predictors of pathologic Q_Alb_ levels within CAA patients. Abbreviations: AD, Alzheimers disease; BG PVS, basal ganglia perivascular spaces; CAA, cerebral amyloid angiopathy; CSVD, cerebral small vessel disease; cSS, cortical superficial siderosis; CSO PVS, centrum semiovale perivascular spaces; MB, microbleeds; MRI, magnetic resonance imaging; WMH, white matter hyperintensities.

|  | All CAA patients<br>n = 115 | Pathologic Q<sub>Alb</sub><br>(Reiber)<br>n = 46 | Remainder of cohort<br>n = 69 | p-value |
| --- | --- | --- | --- | --- |
| **Demographics** |  |  |  |  |
| Age | 76 (71–81) | 79 (72–83) | 75 (70–80) | 0.868 |
| Female sex | 46 (40.0%) | 13 (28.3%) | 33 (47.8%) | **0.036** |
| **Clinical presentation** |  |  |  |  |
| Cognitive impairment | 73 (63.5%) | 28 (60.9%) | 45 (65.2%) | 0.635 |
| Hemorrhagic onset | 40 (34.8%) | 19 (41.3%) | 21 (30.4%) | 0.231 |
| Prior lobar intracerebral hemorrhage | 32 (27.8%) | 15 (32.6%) | 17 (24.6%) | 0.350 |
| Convexity subarachnoid hemorrhage | 13 (11.3%) | 8 (17.4%) | 5 (7.2%) | 0.092 |
| Transient focal neurological episodes | 12 (10.4%) | 7 (15.2%) | 5 (7.2%) | 0.171 |
| Prior ischemic stroke | 31 (27.0%) | 9 (19.6%) | 22 (31.9%) | 0.145 |
| Prior seizure | 28 (24.3%) | 14 (30.4%) | 14 (20.3%) | 0.214 |
| **Vascular risk factors** |  |  |  |  |
| Hypertension | 99 (86.1%) | 38 (82.6%) | 61 (88.4%) | 0.379 |
| Dyslipidemia | 43 (37.4%) | 14 (30.4%) | 29 (42.0%) | 0.208 |
| Diabetes | 36 (31.3%) | 18 (39.1%) | 18 (26.1%) | 0.139 |
| **CSVD markers on MRI** |  |  |  |  |
| Focal cSS presence | 12 (10.4%) | 4 (8.7%) | 8 (11.6%) | 0.618 |
| Disseminated cSS presence | 18 (15.7%) | 12 (26.1%) | 6 (8.7%) | **0.012** |
| Number of lobar MB | 5 (2–21) | 6 (2–29) | 5 (2–17) | 0.602 |
| >10 lobar MB | 43 (37.4%) | 18 (39.1%) | 25 (36.2%) | 0.753 |
| WMH Fazekas scale [0–6] | 5 (4–6) | 5 (4–6) | 5 (4–6) | 0.563 |
| Multiple WMH spots (>10) | 82 (73.9%) | 34 (73.9%) | 48 (73.8%) | 0.994 |
| Peri-basal ganglia WMH | 11 (9.9%) | 5 (10.9%) | 6 (9.2%) | 0.776 |
| Posterior WMH patches | 75 (67.6%) | 30 (65.2%) | 45 (69.2%) | 0.656 |
| Anterior WMH patches | 24 (21.6%) | 8 (17.4%) | 16 (24.6%) | 0.362 |
| Presence of lobar lacune | 35 (30.4%) | 11 (23.9%) | 24 (34.8%) | 0.215 |
| Presence of deep lacune | 25 (21.7%) | 11 (23.9%) | 14 (20.3%) | 0.644 |
| High-degree CSO PVS | 105 (93.8%) | 42 (91.3%) | 63 (95.5%) | 0.372 |
| High-degree BG PVS | 33 (29.5%) | 10 (21.7%) | 23 (34.8%) | 0.134 |
| **Neurodegenerative co-pathologies** |  |  |  |  |
| Amyloid pathology (A+) | 47 (40.9%) | 22 (47.8%) | 25 (36.2%) | 0.215 |
| AD pathology (A+T+) | 28 (24.3%) | 13 (28.3%) | 15 (21.7%) | 0.425 |

**Table 4.** Independent associations between disseminated cortical superficial siderosis and pathologic Q_Alb_ in CAA patients. Abbreviations: cSS, cortical superficial siderosis.

| Dependant variable | Independent variables | OR [95% CI] | p-value |
| --- | --- | --- | --- |
| Pathologic Q<sub>Alb</sub><br>(Reiber) | Age | 0.99 [0.94–1.05] | 0.800 |
|  | Female sex | 0.42 [0.18–0.95] | **0.038** |
|  | Disseminated cSS | 3.92 [1.31–11.72] | **0.014** |

## Discussion

In this study, we demonstrate that Q_Alb_ is elevated in patients with probable CAA compared with AD patients and cognitively unimpaired controls. This association remained significant after adjustment for demographic variables and vascular risk factors, suggesting that increased Q_Alb_ reflects disease-related barrier dysfunction rather than differences in vascular comorbidity alone. Importantly, Q_Alb_ showed only low-to-moderate diagnostic accuracy for distinguishing CAA from comparison groups, arguing against a stand-alone CAA-specific diagnostic biomarker. However, the most pathophysiologically relevant finding was observed within the CAA cohort: pathological Q_Alb_ levels were independently associated with disseminated cSS, a hemorrhagic marker linked to advanced leptomeningeal vascular injury and incident ICH risk. Together, these findings support the concept that Q_Alb_ may capture a biologically meaningful barrier-dysfunction phenotype within CAA.

Albumin is synthesized in the periphery and enters the CSF only in small amounts under physiological conditions [22]. Therefore, Q_Alb_ is widely used as an accessible marker of blood–CSF barrier integrity and global barrier permeability [23]. A recent multimodal study combined contrast-enhanced MRI with CSF analyses demonstrating that elevated Q_Alb_ is associated with inflammatory BBB dysfunction on neuroimaging, and associated with increased concentrations of inflammatory cytokines (IL-6, IL-8, TNF-α), matrix metalloproteinases (MMP-2, MMP-3, MMP-10), and angiogenic factors [24].

Our findings fit well into current pathophysiological frameworks of CAA. It is increasingly recognized as a dynamic neurovascular disease evolving from vascular amyloid accumulation to impaired vascular function, barrier dysfunction, tissue injury, and ultimately hemorrhagic complications [25]. In this framework, BBB dysfunction may represent an intermediate disease process, linking vascular amyloid pathology to downstream hemorrhagic and inflammatory manifestations. The observation that Q_Alb_ was elevated in CAA compared with AD is therefore notable, because both disorders share amyloid-related biology, but CAA is distinguished by primary involvement of leptomeningeal and cortical vessels. Thus, increased Q_Alb_ may indicate the vascular component of amyloid disease rather than amyloid pathology per se. The attenuated RoM effect size in AD compared to the effect size in comparison with controls supports that hypothesis.

Several imaging studies support the presence of barrier dysfunction in CAA. Contrast-enhanced MRI studies have demonstrated BBB leakage in patients with probable CAA, including leakage at the level of leptomeningeal and small parenchymal vessels [3, 5, 26, 27]. Of note, the severity of contrast extravasation in CAA was associated with cSS severity and Q_Alb_ [5, 27]. Other work has suggested that subarachnoid CSF hyperintensities at MRI may reflect subtle leakage of blood proteins or products into the CSF compartment [25, 28]. These imaging observations are highly complementary to our CSF findings. While contrast-enhanced MRI provides spatial information on regional leakage, Q_Alb_ may capture a more global signal of barrier dysfunction.

The association between pathological Q_Alb_ and disseminated cSS within CAA patients is a key finding of this study. cSS is considered a chronic manifestation of repeated bleeding into the superficial cortical or subarachnoid compartment and is among the strongest imaging predictors of incident lobar ICH in CAA [29, 30]. Neuropathological studies indicate that cSS corresponds to iron-positive deposits in superficial cortical layers and is particularly linked to advanced leptomeningeal CAA-related vessel wall damage [29]. Our finding that disseminated cSS, but not the burden of lobar microbleeds or white matter hyperintensities, was independently associated with pathological Q_Alb_ suggests that barrier dysfunction may be especially relevant in the leptomeningeal–subarachnoid hemorrhagic phenotype of CAA. This supports the hypothesis that Q_Alb_ may identify a subgroup of patients with more active, leaky, and hemorrhage-prone vascular disease.

The link between cSS and Q_Alb_ may also have an inflammatory component. cSS is not merely an inert residue of previous bleeding; experimental and neuropathological data suggest that superficial iron deposition is associated with reactive astrogliosis and local neuroinflammatory responses [6, 31]. In parallel, postmortem studies in CAA have identified perivascular inflammation and vascular remodeling around vulnerable vessels, raising the possibility that barrier disruption, leakage of blood-derived proteins, iron deposition, and inflammatory activation may reinforce each other [32]. In this model, elevated Q_Alb_ could either precede hemorrhagic leakage by indicating vascular fragility or follow repeated superficial bleeding as a marker of secondary inflammatory barrier damage. Because our study is cross-sectional, these temporal relationships cannot be resolved, but the association with disseminated cSS strongly supports further longitudinal investigation.

These observations also resonate with the emerging concept of CAA-related inflammation spectrum disorders. Classical CAA-related inflammation represents the clinically overt end of this spectrum, but recent work suggests that more subtle inflammatory manifestations may occur in patients who do not fulfill established CAA-ri criteria [4, 33]. This is particularly relevant for patients with cSS, convexity subarachnoid hemorrhage, and transient focal neurological episodes, in whom meningeal or vessel-wall enhancement and inflammatory CSF changes may be present despite incomplete CAA-ri phenotypes. In this context, Q_Alb_ should not be considered a diagnostic marker for CAA-ri, but it may represent a low-threshold fluid marker of barrier dysfunction that could help identify patients with a more inflammatory or biologically active CAA phenotype.

An important aspect of our study is the comparison with AD patients. BBB dysfunction has been described in AD and other dementias, but the degree and biological context of barrier impairment may differ substantially between neurodegenerative and vascular disorders. In our cohort, Q_Alb_ was higher in CAA than in AD despite the overlap between both diseases in amyloid biology and age-related comorbidity. This suggests that Q_Alb_ may capture the vascular injury component of CAA. Nevertheless, the observed overlap between groups and the modest AUC values indicate that Q_Alb_ lacks sufficient specificity for diagnostic classification. Its value may therefore be more likely pathophysiological and prognostic than diagnostic. This interpretation is also consistent with previous studies of core CSF biomarkers in CAA. Classical AD-related CSF markers, including Aβ40, Aβ42, tau, and phosphorylated tau, provide information on amyloid and neurodegenerative co-pathology but have limited diagnostic utility for CAA [34, 35, 8]. Q_Alb_ addresses a further biological axis: barrier integrity. Therefore, Q_Alb_ should not be viewed as a competitor to amyloid or tau markers, but as a complementary marker that may capture vascular permeability and inflammatory barrier dysfunction. Future biomarker panels in CAA may need to combine amyloid-related markers, neurodegeneration markers, inflammatory mediators, and barrier markers to reflect the full biological heterogeneity of the disease.

Clinically, our findings do not support Q_Alb_ as a stand-alone diagnostic test for CAA. However, Q_Alb_ is inexpensive, widely available in routine CSF diagnostics, and biologically interpretable. Its potential relevance may lie in risk phenotyping rather than disease detection. Patients with elevated Q_Alb_ and disseminated cSS may represent a subgroup with more advanced leptomeningeal vessel injury, greater barrier dysfunction, and possibly higher hemorrhagic activity. If confirmed longitudinally, Q_Alb_ could help enrich cohorts for studies on hemorrhage prediction, CAA progression, or anti-inflammatory and vasoprotective treatment strategies. At present, this remains hypothesis-generating and should not be translated into individual risk prediction without prospective validation.

### Limitations

Several limitations need to be acknowledged. First, this was a retrospective, single-center study, and selection bias cannot be excluded. The dependence on CAA patients who underwent lumbar puncture and CSF analyses might enriched for diagnostically complex cases. Second, controls were not population-based healthy controls but memory-clinic participants without cognitive impairment and without hemorrhagic MRI markers, which may limit generalizability. Third, the study groups differed in demographics and vascular risk factors. Although we adjusted for these variables, residual confounding cannot be excluded. Fourth, Q_Alb_ is a global marker and does not provide spatial information on the location of barrier dysfunction. Finally, the cross-sectional design precludes conclusions about causality or temporal sequence; we cannot determine whether increased Q_Alb_ precedes cSS, follows repeated superficial bleeding, or reflects a shared underlying inflammatory vascular process.

### Future directions

Future studies should validate these findings in multicenter, prospectively recruited cohorts. Longitudinal designs are required to determine whether Q_Alb_ predicts incident hemorrhage, progression of cSS or development of CAA-related inflammation. Multimodal approaches combining Q_Alb_ with contrast-enhanced MRI, and inflammatory mediators may help to disentangle the relationship between barrier dysfunction, neuroinflammation, and hemorrhagic progression. In summary, elevated Q_Alb_ in CAA, particularly its association with disseminated cSS, supports the concept that barrier dysfunction is closely linked to the hemorrhage-prone phenotype of CAA and may provide a clinically accessible window into vascular integrity and inflammatory disease activity in CAA.

## Acknowledgments

This study received no funding.

## Author Contributions

P.A. and S.S. contributed to the conception and design of the study; B.B., P.A., C.G., and M.P. contributed to the acquisition and analysis of data; all authors contributed to drafting the text.

## Data Availability

The data that support the findings of this study are avail-able from the corresponding author on reasonable reques

## References

1. Koemans EA, Chhatwal JP, van Veluw SJ, et al. (2023) Progression of cerebral amyloid angiopathy: a pathophysiological framework. Lancet Neurol 22:632–642. 10.1016/S1474-4422(23)00114-X

2. Raposo N, Morice Morand M, Genin T et al. (2026) Cerebral Amyloid Angiopathy and Risk of Dementia in Patients With Cognitive Complaint. Neurology 106:e218009. 10.1212/WNL.0000000000218009

3. Sellimi A, Panteleienko L, Mallon D et al. (2025) Inflammation in Cerebral Amyloid Angiopathy-Related Transient Focal Neurological Episodes. Ann Neurol 97:475–482. 10.1002/ana.27164

4. Charidimou A (2025) Cerebral Amyloid Angiopathy-Related Inflammation Spectrum Disorders: Introduction of a Novel Concept and Diagnostic Criteria. Ann Neurol 97:470–474. 10.1002/ana.27162

5. van den Brink H, Kozberg MG, Makkinejad N, et al. (2025) In Vivo Imaging of Blood-Brain Barrier Leakage Using a Contrast Agent in Patients With Cerebral Amyloid Angiopathy: An Exploratory Study. Neurology 105:e214336. 10.1212/WNL.0000000000214336

6. van den Brink H, Voigt S, Kozberg M, et al. (2024) The role of neuroinflammation in cerebral amyloid angiopathy. EBioMedicine 110:105466. 10.1016/j.ebiom.2024.105466

7. Varma C, Lemere CA (2025) CAA proteomics meta-analysis reveals novel targets, key players, and the effects of sex, APOE, and brain region in humans. Acta Neuropathol 149:40. 10.1007/s00401-025-02886-3

8. Arndt P, Pfister M, Perosa V et al. (2025) Risk factors and clinical significance of neurodegenerative co-pathologies in symptomatic cerebral small vessel disease. J Neurol 272:349. 10.1007/s00415-025-13087-z

9. Charidimou A, Boulouis G, Frosch MP et al. (2022) The Boston criteria version 2.0 for cerebral amyloid angiopathy: a multicentre, retrospective, MRI-neuropathology diagnostic accuracy study. Lancet Neurol 21:714–725. 10.1016/S1474-4422(22)00208-3

10. McKhann G, Drachman D, Folstein M et al. Clinical diagnosis of Alzheimer’s disease: Report of Department of Health and Human Services Task Force on Alzheimer’s Disease:939–944

11. Jack CR, Jr., Bennett DA, Blennow K et al. (2018) NIA-AA Research Framework: Toward a biological definition of Alzheimer’s disease. Alzheimers Dement 14:535–562. 10.1016/j.jalz.2018.02.018

12. Hußler W, Höhn L, Stolz C et al. (2022) Brevican and Neurocan Cleavage Products in the Cerebrospinal Fluid - Differential Occurrence in ALS, Epilepsy and Small Vessel Disease. Front Cell Neurosci 16:838432. 10.3389/fncel.2022.838432

13. Wardlaw JM, Smith EE, Biessels GJ et al. (2013) Neuroimaging standards for research into small vessel disease and its contribution to ageing and neurodegeneration. Lancet Neurol 12:822–838. 10.1016/S1474-4422(13)70124-8

14. Duering M, Biessels GJ, Brodtmann A et al. (2023) Neuroimaging standards for research into small vessel disease-advances since 2013. Lancet Neurol 22:602–618. 10.1016/S1474-4422(23)00131-X

15. Gregoire SM, Chaudhary UJ, Brown MM et al. (2009) The Microbleed Anatomical Rating Scale (MARS): reliability of a tool to map brain microbleeds. Neurology 73:1759–1766. 10.1212/WNL.0b013e3181c34a7d

16. Charidimou A, Schmitt A, Wilson D et al. (2017) The Cerebral Haemorrhage Anatomical RaTing inStrument (CHARTS): Development and assessment of reliability. J Neurol Sci 372:178–183. 10.1016/j.jns.2016.11.021

17. Wahlund LO, Barkhof F, Fazekas F et al. A New Rating Scale for Age-Related White Matter Changes Applicable to MRI and CT

18. Charidimou A, Boulouis G, Haley K et al. (2016) White matter hyperintensity patterns in cerebral amyloid angiopathy and hypertensive arteriopathy. Neurology 86:505–511. 10.1212/WNL.0000000000002362

19. Friedrich JO, Adhikari NKJ, Beyene J (2008) The ratio of means method as an alternative to mean differences for analyzing continuous outcome variables in meta-analysis: a simulation study. BMC Med Res Methodol 8:32. 10.1186/1471-2288-8-32

20. Friedrich JO, Adhikari NKJ, Beyene J (2012) Ratio of geometric means to analyze continuous outcomes in meta-analysis: comparison to mean differences and ratio of arithmetic means using empiric data and simulation. Stat Med 31:1857–1886. 10.1002/sim.4501

21. Reiber H Cerebrospinal fluid: Laboratory Analysis, Evaluation and Interpretation, 2020th edn.

22. Reiber H (2001) Dynamics of brain-derived proteins in cerebrospinal fluid. Clinica Chimica Acta 310:173–186

23. Andersson M, Alvarez-Cermeño J, Bernardi G et al. (1994) Cerebrospinal fluid in the diagnosis of multiple sclerosis: a consensus report. Journal of Neurology, Neurosurgery and Psychiatry 57:897–902

24. Hillmer L, Erhardt EB, Caprihan A et al. (2023) Blood-brain barrier disruption measured by albumin index correlates with inflammatory fluid biomarkers. J Cereb Blood Flow Metab 43:712–721. 10.1177/0271678X221146127

25. Koemans EA, van Walderveen MAA, Voigt S, et al. (2023) Subarachnoid CSF hyperintensities at 7 tesla FLAIR MRI: A novel marker in cerebral amyloid angiopathy. Neuroimage Clin 38:103386. 10.1016/j.nicl.2023.103386

26. Voigt S, Jong J de, Voorter P, et al. (2026) Contrasting patterns of leptomeningeal and parenchymal gadolinium extravasation in cerebral amyloid angiopathy: An MRI-based evaluation. Int J Stroke:17474930261424089. 10.1177/17474930261424089

27. Arndt P, Khadhraoui E, Müller SJ et al. (2026) Meningovascular Inflammation in Cerebral Amyloid Angiopathy-Related Cortical Superficial Siderosis. Ann Clin Transl Neurol. 10.1002/acn3.70315

28. Panteleienko L, Banerjee G, Mallon DH et al. (2024) Sulcal Hyperintensity as an Early Imaging Finding in Cerebral Amyloid Angiopathy-Related Inflammation. Neurology 103:e210084. 10.1212/WNL.0000000000210084

29. Charidimou A, Perosa V, Frosch MP et al. (2020) Neuropathological correlates of cortical superficial siderosis in cerebral amyloid angiopathy. Brain 143:3343–3351. 10.1093/brain/awaa266

30. Charidimou A, Boulouis G, Greenberg SM et al. (2019) Cortical superficial siderosis and bleeding risk in cerebral amyloid angiopathy: A meta-analysis. Neurology 93:e2192–e2202. 10.1212/WNL.0000000000008590

31. Auger CA, Perosa V, Greenberg SM et al. (2023) Cortical superficial siderosis is associated with reactive astrogliosis in cerebral amyloid angiopathy. J Neuroinflammation 20:195. 10.1186/s12974-023-02872-0

32. Kozberg MG, Yi I, Freeze WM et al. (2022) Blood-brain barrier leakage and perivascular inflammation in cerebral amyloid angiopathy. Brain Commun 4:fcac245. 10.1093/braincomms/fcac245

33. Banerjee G, Werring DJ (2025) Hiding in Plain Sight: Inflammation in Iatrogenic Cerebral Amyloid Angiopathy. Neurol Neuroimmunol Neuroinflamm 12:e200493. 10.1212/NXI.0000000000200493

34. Charidimou A, Boulouis G (2024) Core CSF Biomarker Profile in Cerebral Amyloid Angiopathy: Updated Meta-Analysis. Neurology 103:e209795. 10.1212/WNL.0000000000209795

35. Margraf NG, Jensen-Kondering U, Weiler C et al. (2022) Cerebrospinal Fluid Biomarkers in Cerebral Amyloid Angiopathy: New Data and Quantitative Meta-Analysis. Front Aging Neurosci 14:783996. 10.3389/fnagi.2022.783996

